# Study protocol for a randomized clinical trial evaluating the effectiveness of a Brazilian lifestyle-based diabetes prevention program: the PROVEN-DIA trial

**DOI:** 10.64898/2026.07.11.26357806

**Authors:** Angela Cristine Bersch-Ferreira, Raira Pagano, Danielle Cristina Fonseca, Thatiane Lopes Valentim Di Paschoale Ostolin, Ana Laura Fogaça, Livia Tavares De Oliveira, Angelica Barbosa Neres Santana, Barbara Shibuya Alves, Cleyton Zanardo De Oliveira, Aline Marcadenti De Oliveira, Ana Paula Perillo Ferreira Carvalho, Ana Teresa Mana Gonçalves Santomauro, Augusto Cezar Santomauro, Bernardete Weber, Enilda Maria de Sousa Lara, Josefina Bressan, Jussara Carnevale de Almeida, Marcelo Macedo Rogero, Sônia Lopes Pinto, Viviane Sahade, Bianca de Almeida-Pititto, Daniela Lopes Gomes, Dhiãnah Santini De Oliveira Chachamovitz, Adriano Namo Cury

**Author notes:** These authors contributed equally to this work (Angela Cristine Bersch-Ferreira, Raira Pagano, Danielle Cristina Fonseca, Thatiane Lopes Valentim Di Paschoale Ostolin, Ana Laura Fogaça, Livia Tavares De Oliveira, Angelica Barbosa Neres Santana, Barbara Shibuya Alves, Cleyton Zanardo De Oliveira). These authors also contributed equally to this work (Aline Marcadenti De Oliveira, Ana Paula Perillo Ferreira Carvalho, Ana Teresa Mana Gonçalves Santomauro, Augusto Cezar Santomauro, Bernardete Weber, Enilda Maria de Sousa Lara, Josefina Bressan, Jussara Carnevale de Almeida, Marcelo Macedo Rogero, Sônia Lopes Pinto, Viviane Sahade, Bianca de Almeida-Pititto, Daniela Lopes Gomes, Dhiãnah Santini De Oliveira Chachamovitz, Adriano Namo Cury).

## Abstract

Type 2 diabetes (T2D) affects more than 16 million Brazilians and has nearly doubled in the last 20 years. Lifestyle interventions reduce T2D incidence in people at risk of developing the disease; however, no large-scale trials have evaluated diabetes prevention programs in Brazil or compared telehealth and hybrid delivery in middle-income settings. These are key gaps that must be addressed to enable nationwide scale-up, particularly in primary care settings and remote areas. This article presents the protocol for the PROVEN-DIA trial, designed to address these evidence gaps. PROVEN-DIA is a multicenter, open-label, randomized controlled superiority trial with a parallel design enrolling 1,305 adults with prediabetes at 30 sites across all five Brazilian regions (ClinicalTrials.gov: NCT06426277). Participants will be randomly assigned in equal numbers to one of three groups. All groups receive lifestyle guidance targeting diet, physical activity, sleep, stress, alcohol consumption, and smoking, in accordance with Brazilian national guidelines. The two intervention groups receive PROVEN-DIA, a structured 36-month lifestyle program with 43 scheduled contacts encompassing individual counseling sessions, structured support contacts, and group education sessions, delivered either in a hybrid format (PROVEN-DIA) or telehealth only (TelePROVEN-DIA). The control group receives the same lifestyle guidance through unstructured individual visits every six months, without predefined content or ongoing support. The primary outcome is the cumulative incidence of type 2 diabetes at 36 months. Secondary outcomes include body weight, fasting glucose, glycated hemoglobin (HbA1c), dietary quality, physical activity, sedentary behavior, sleep quality, perceived stress, alcohol consumption, smoking behavior, and health-related quality of life. Analyses will follow the intention-to-treat principle.

## Introduction

Type 2 diabetes (T2D) is a major global health challenge, with prevalence projected to increase from 589 million adults in 2024 to 853 million by 2050 [1]. Brazil ranks sixth worldwide, with prevalence rising from 5.5% in 2006 to 10.2% in 2023 [1,2]. In low- and middle-income countries, where most new diagnoses occur, substantial gaps persist in access to treatment and disease control [3,4]. Prediabetes represents a critical window for intervention to prevent new cases. In Brazil, the prevalence of prediabetes is approximately 18.5% among adults, highlighting a substantial at-risk population and the need for effective prevention strategies [5]. Strong evidence from randomized trials shows that structured lifestyle modification programs can cut the risk of progressing to T2D by up to 58% over an average of 3 years with sustained long-term benefits, as demonstrated in landmark studies including the US Diabetes Prevention Program (DPP) and the Finnish Diabetes Prevention Study (DPS), China Da Qing Study, and Indian Diabetes Prevention Program [6–9].

These interventions were grounded in healthy habits, particularly nutrition, physical activity, and sleep, as primary behavioral targets for preventing chronic diseases, with additional lifestyle factors such as stress management and substance use potentially contributing to risk reduction [10,11]. Despite this evidence, critical implementation gaps remain in Brazil. First, structured diabetes prevention programs have not been implemented at scale: generalizability from trials conducted in other settings is constrained by differences in healthcare system organization, cultural dietary patterns, and socioeconomic context [12,13], and international programs require substantial cultural adaptation before implementation can be justified [14,15]. Second, substantial heterogeneity exists in prediabetes management across Brazilian primary care, with no standardized, evidence-based protocol currently available [16]. Third, Brazil’s vast geographic dimensions pose unique implementation challenges, as access to healthcare is severely restricted in remote areas due to large distances, inadequate infrastructure, and economic barriers [17,18]. Although telehealth has emerged as a promising strategy to overcome these barriers [19,20], its effectiveness for diabetes prevention compared to hybrid delivery has not been rigorously evaluated in this context.

Considering these gaps, the PROVEN-DIA program (PROgrama de preVENção de DIAbetes) was designed as a culturally tailored lifestyle intervention aligned with Brazilian national guidelines, validated through expert consensus and pilot-tested in a multicenter trial demonstrating feasibility [21,22]. This article presents the protocol for a multicenter randomized controlled trial evaluating whether PROVEN-DIA is more effective than usual care in preventing T2D among Brazilian adults with prediabetes, comparing two scalable delivery formats: hybrid (in-person plus remote) and telehealth-only.

## Materials and Methods

### Study Design

The primary objective of the PROVEN-DIA trial is to evaluate the effectiveness of a structured lifestyle-based diabetes prevention program in reducing the incidence of T2D among Brazilian adults with prediabetes, comparing two delivery formats [hybrid, a mix of in-person and remote support (PROVEN-DIA), and telehealth-only (TelePROVEN-DIA)] against usual care. Secondary objectives include assessing intervention effects on body weight, fasting glucose and glycated hemoglobin (HbA1c) levels, dietary quality, physical activity, sedentary behavior, sleep quality, perceived stress, alcohol consumption, smoking behavior, and health-related quality of life.

This is a multicenter, randomized, controlled, open-label, superiority trial with three parallel groups (1:1:1 allocation ratio) and 36-month follow-up. Hospital BP (A Beneficência Portuguesa de São Paulo) serves as the coordinating center, and the trial is conducted across 30 sites in all five Brazilian regions. The Coordinating Center is responsible for trial management, data integrity, statistical oversight, and quality assurance. The Steering Committee, composed of investigators from pilot study sites and external independent researchers, provides scientific oversight and strategic guidance on protocol implementation, trial conduct, and interpretation of findings. Participating research sites were selected based on the investigators’ experience in conducting lifestyle intervention studies aimed at the prevention or management of chronic conditions, or on their previous successful collaboration with the coordinating center in the implementation of lifestyle modification clinical trials. This strategy sought to ensure the operational feasibility of the study while maintaining geographic diversity and representation across different regions of Brazil.

### Trial registration and protocol approval

The PROVEN-DIA trial protocol was developed in accordance with the SPIRIT 2025 Statement (S1 Appendix) and registered in ClinicalTrials.gov (Identifier: NCT06426277). The protocol was approved by the Research Ethics Committee of Hospital BP (Certificate of Presentation for Ethical Consideration – CAAE: 75963123.8.0000.5483) as the central ethics committee and subsequently approved by the local Research Ethics Committees of all investigation sites prior to the initiation of recruitment at each site. The trial is conducted in accordance with the principles of the Declaration of Helsinki, Brazilian National Health Council Resolution No. 466/2012, the Brazilian Clinical Research Law (Law No. 14,874/2024), the International Council for Harmonization Good Clinical Practice (ICH-GCP) guidelines, and the World Health Organization (WHO) ethical principles. Written informed consent is obtained by trained investigators from all eligible individuals prior to any study-related procedures. Any protocol amendments are formally documented, updated in the trial registry, and communicated to all relevant stakeholders, including ethics committees, investigation sites, and, when applicable, study participants.

### Study Population

Participants are recruited at investigation sites through institutional campaigns, active database searches, and community announcements. Interested individuals complete a standardized pre-screening form, which includes the Brazilian Portuguese version of the Finnish Diabetes Risk Score [23,24] and a preliminary assessment of key eligibility criteria. Those identified as potentially eligible are invited to attend the investigation site for a detailed study presentation and formal eligibility confirmation. Individuals who do not meet eligibility criteria at pre-screening are not enrolled but receive brief guidance on healthy eating and physical activity based on Brazilian national guidelines, ensuring that participation in the pre-screening process is beneficial regardless of eligibility.

Eligible participants are adults aged ≥18 years with a Body Mass Index (BMI) between 18.5 and 34.9 kg/m², who have access to an electronic device (computer, tablet, or smartphone) with internet connectivity and can access the investigation site within 60 minutes. Participants must meet the study definition of prediabetes, confirmed through two HbA1c measurements between 5.7% and 6.4% collected 7 to 90 days apart [25]. Participants must not have received individualized nutritional counseling or individualized supervised physical training within the previous six months.

Key exclusion criteria include a confirmed diagnosis of diabetes mellitus, recent or current use of glucose-lowering medications (within three months), or medical conditions likely to significantly limit life expectancy or interfere with study participation. Further exclusion criteria encompass conditions or behaviors likely to affect program adherence, including current pregnancy or breastfeeding, severe psychiatric disorders precluding participation at the clinical team’s discretion, excessive alcohol consumption, current or recent participation in another clinical trial (within six months), anticipated relocation outside the study area during follow-up, and household membership with a study participant or PROVEN-DIA team member, or prior participation in the pilot study. Current use of systemic corticosteroids, antineoplastic agents, psychoactive agents, selective serotonin reuptake inhibitors, or weight loss medications is also exclusionary. A complete list of eligibility criteria is provided in S2 Appendix.

### Randomization and Blinding

The allocation sequence is generated centrally by the Coordinating Center using computer-based randomization with scripts developed in R. Randomization uses variable block sizes (3, 6, or 9), stratified by investigation sites to ensure balanced allocation across centers, with a 1:1:1 ratio. Allocation concealment is maintained through a secure, password-protected, web-based platform (REDCap - https://redcap.bp.org.br) available 24 hours a day. The REDCap Randomization module performs participant allocation and reveals group assignment only after participant registration is complete and all eligibility criteria are confirmed. Investigators and site staff do not have access to the randomization sequence and cannot predict allocation.

Due to the nature of the lifestyle intervention, the trial is open-label, neither participants nor intervention providers can be blinded to group assignment. To minimize potential contamination, participants from different trial arms are scheduled for in-person assessments on separate days. Participant sessions are conducted exclusively within each trial arm, with no cross-arm contact planned during group activities, and participants are advised at enrollment to avoid sharing intervention-specific content with individuals from other groups. Outcome assessors responsible for laboratory measurements are blinded to group allocation, and statisticians remain blinded to intervention assignments throughout the analysis phase.

### Interventions

The study includes three parallel arms: PROVEN-DIA, delivering the structured program in a hybrid format (in-person and remote); TelePROVEN-DIA, delivering the same structured program exclusively via telehealth; and a control group receiving usual care. Across all arms, the overarching objective is to promote lifestyle changes associated with diabetes prevention, including improving diet quality with reduced consumption of ultra-processed foods, achieving at least 150 minutes per week of moderate-to-vigorous physical activity, and fostering healthier sleep, stress management, alcohol consumption, and smoking behaviors, in accordance with Brazilian Dietary and Physical Activity Guidelines [26,27]. To support adherence across all three groups, participants receive lifestyle change materials (a thermal water bottle, a T-shirt, and a cap) provided as engagement incentives. The intervention was designed to be delivered by healthcare professionals from different disciplines, reflecting the multidisciplinary workforce typically available in primary healthcare settings within the Brazilian Unified Health System (SUS). Professionals from fields such as medicine, nursing, nutrition, physical education, physiotherapy, pharmacy, and other health-related disciplines were eligible to deliver the intervention. No intervention component involved activities restricted to a specific professional category under Brazilian regulations. This approach was intentionally adopted to enhance the feasibility, scalability, and future implementation of the program in real-world primary care settings.

Unlike the control group, the PROVEN-DIA and TelePROVEN-DIA arms deliver lifestyle guidance through a standardized, structured program culturally adapted to the Brazilian context. The program applies the Transtheoretical Model of Behavior Change [28] to guide participants through readiness-based SMART goal-setting (Specific, Measurable, Achievable, Relevant, Time-bound) for each targeted lifestyle behavior [29,30]. To support adherence and self-awareness, participants are provided with self-monitoring tools to track dietary intake, physical activity, sleep quality, stress levels, alcohol consumption, and smoking behavior. These tools allow participants to monitor their own progress and identify behavioral patterns, with data reviewed during counseling sessions to reinforce goal-setting and behavioral planning. The program was developed and validated using the Delphi technique with a multidisciplinary expert panel [21]. To facilitate transparency and replication, the PROVEN-DIA intervention was described according to the Template for Intervention Description and Replication (TIDieR) Checklist as provided in S3 Appendix. Both arms follow identical interaction structures and frequencies, comprising 43 scheduled contacts over 36 months (Fig S4.2), differing only in delivery mode. The interactions include data collection visits, one-on-one counseling sessions with a Lifestyle Counselor, structured follow-up contacts with a Lifestyle Coach, and group education sessions (Table 1). Lifestyle Counselors are trained healthcare professionals responsible for individualized behavioral counseling, readiness-to-change assessment, instruction on self-monitoring tools, and collaborative goal-setting. Lifestyle Coaches provide structured support between counseling sessions to reinforce behavioral goals and adherence to self-monitoring practices. In the PROVEN-DIA arm, Lifestyle Counselor consultations occur in-person while Lifestyle Coach contacts are conducted remotely; in the TelePROVEN-DIA arm, all interactions are delivered remotely via videoconferencing and centrally coordinated. A detailed description of the theoretical framework, program phases, and interaction types and frequencies is provided in S4 Appendix.

**Table 1.**
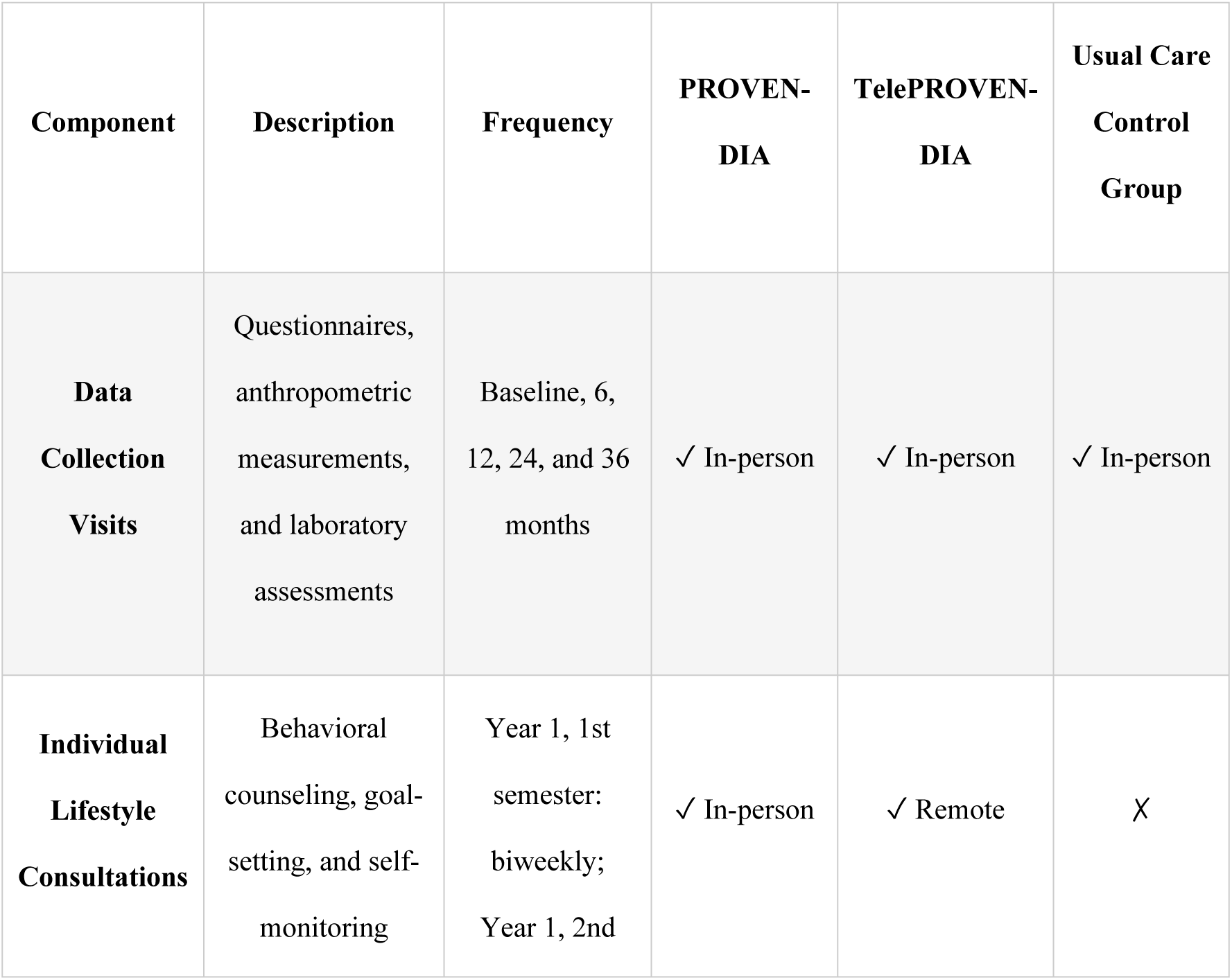
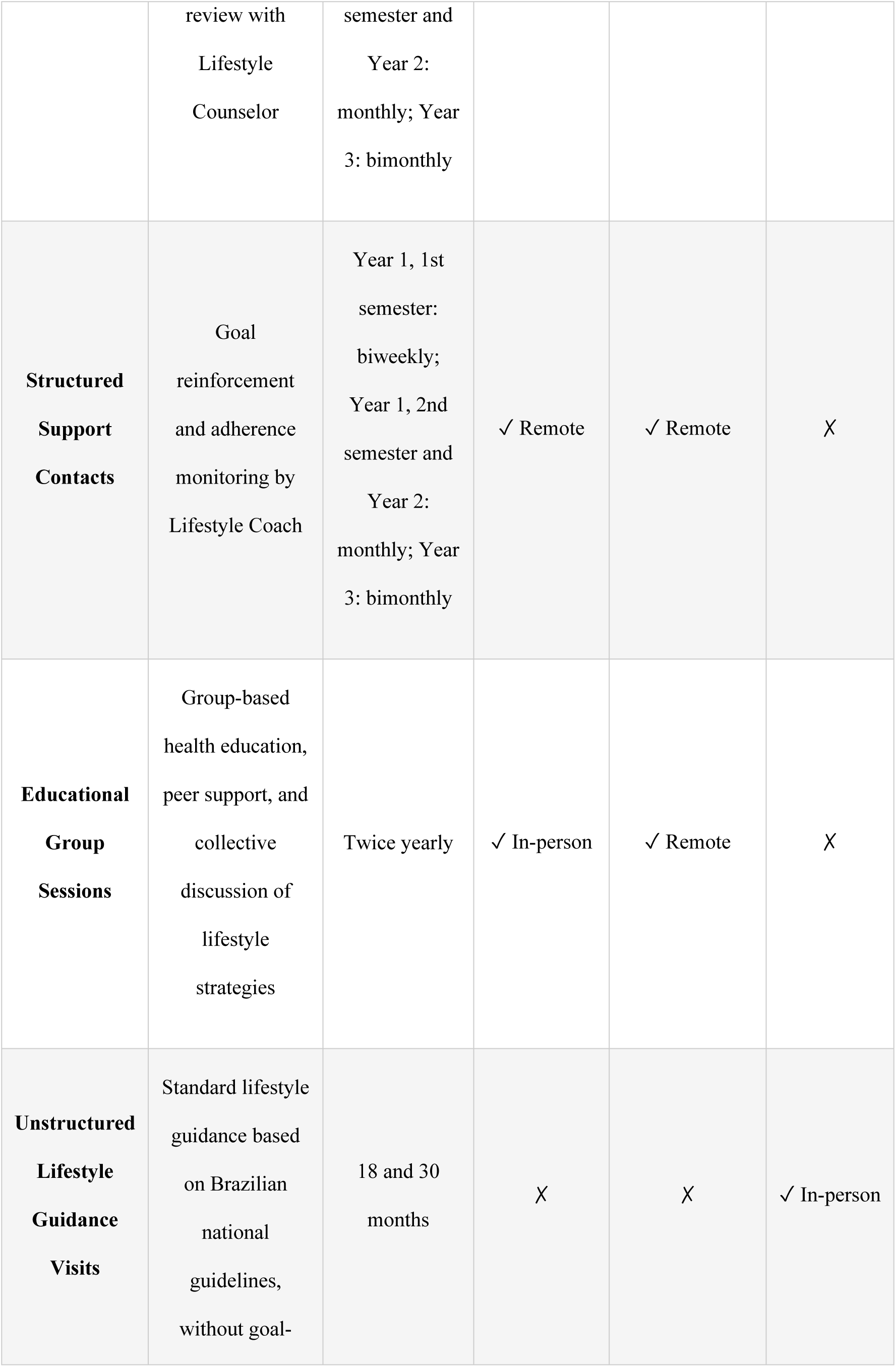
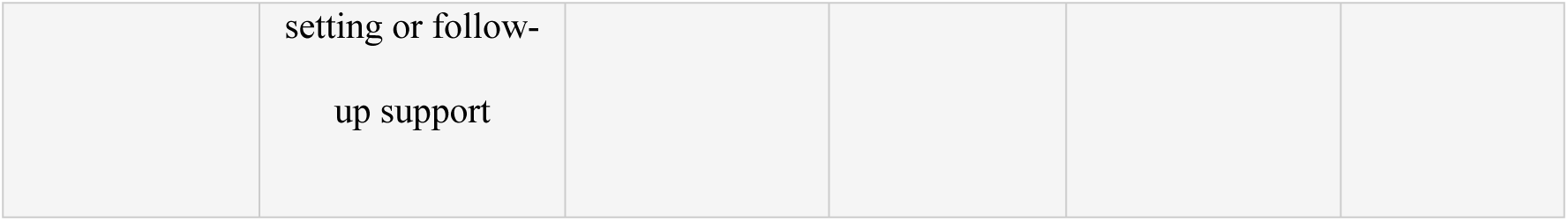
Intervention components by study arm.

Participants allocated to the usual care control group receive the same lifestyle guidance content as the intervention arms (covering diet, physical activity, sleep, stress management, alcohol consumption, and smoking). However, this guidance is not delivered through a structured program: there are no scheduled contacts between visits, no behavioral goal-setting, and no ongoing support. Individual visits are scheduled every six months by trained study staff following a predefined protocol, ensuring consistency across sites. This approach reflects the minimum standard of care recommended for individuals with prediabetes in Brazil while avoiding contamination from heterogeneous local practices. A detailed description of the control group protocol is provided in S5 Appendix.

### Data Collection

At baseline, trained study staff administer a standardized questionnaire to collect sociodemographic information from all participants, including age, sex assigned at birth, gender identity, sexual orientation, self-identified race/ethnicity, presence of physical disability, and marital status. Socioeconomic status and education level are assessed according to the Brazilian Economic Classification Criteria [31].

Clinical assessments comprise medical history, comorbidities, current and past medication use, family history of diabetes, and diabetes risk assessment [23,24]. Although the use of glucose-lowering medications is an exclusion criteria, information on any new use of these medications during follow-up is systematically documented at each visit, as this may influence the interpretation of study outcomes. Additionally, use of dietary supplements and other medications is recorded, including drug type, dosage, frequency, and duration of use.

Anthropometric measurements include weight, BMI, waist and calf circumferences, and vital signs (systolic and diastolic blood pressure and heart rate), all collected using standardized protocols. Laboratory assessments include fasting plasma glucose, fasting insulin, HbA1c, lipid profile (total cholesterol, LDL-cholesterol, non-HDL cholesterol, HDL-cholesterol, and triglycerides), creatinine, urea, and complete blood count. Blood samples are collected after an overnight fast (minimum 8 hours) and analyzed using standardized methods at certified laboratories. Indices of insulin resistance and β-cell function (HOMA-IR and HOMA-β) are calculated from fasting glucose and insulin values [32]. In addition to laboratory analyses, whole blood and plasma samples are collected and stored in a biorepository for future ancillary studies.

Dietary assessment includes food insecurity screening using the Brazilian Food Insecurity Scale (EBIA) [33], dietary intake markers based on the Brazilian Food and Nutrition Surveillance System (SISVAN) [34], food access assessment [35], and two 24-hour dietary recalls.

Physical activity and sedentary behavior are assessed using the long version of the International Physical Activity Questionnaire (IPAQ-long) [36], complemented by a validated instrument assessing environmental factors and perceived social support for physical activity [37].

Psychosocial and behavioral parameters are assessed using instruments validated for the Brazilian population, including perceived stress (Perceived Stress Scale-10, PSS-10) [38], sleep quality (Pittsburgh Sleep Quality Index – Brazilian version, PSQI-BR) [39], health-related quality of life (Short Form-36, SF-36) [40], smoking status and intensity, alcohol consumption frequency and quantity, and mindfulness (Mindful Attention Awareness Scale, MAAS) [41].

Adverse events are systematically documented at all study interactions. In addition, biological samples, including plasma and whole blood (with and without stabilizing additives), are collected and stored at −70 °C for future ancillary analyses.

Data are collected at baseline and at each follow-up timepoint, organized by assessment domain as summarized in Figure 1.

**Fig 1.** Schedule of enrolment, interventions, and assessments (SPIRIT figure). **^a^** Timepoints are expressed in months relative to randomization (month 0). −t₁ denotes the enrolment/screening visit prior to randomization. Data collection visits occur at baseline and at 6, 12, 24, and 36 months (V0–V4). **^b^** Arrow indicates continuous delivery of the structured lifestyle program from randomization to study close-out, organized in four phases of decreasing contact frequency: months 0–6 biweekly, months 6–24 monthly, months 24–36 bimonthly. Contacts comprise Individual Lifestyle Consultations, Structured Support Contacts, and Educational Group Sessions (twice yearly). PROVEN-DIA delivers consultations in person and coach contacts remotely; TelePROVEN-DIA delivers all contacts remotely. See S4 Appendix for the full contact matrix by arm and phase. **^c^** The usual care control group receives unstructured lifestyle guidance at data collection visits (0, 6, 12, 24, 36 months) plus additional standard guidance visits at 18 and 30 months, without goal-setting or ongoing support (S5 Appendix). **^d^** Medical history and comorbidities are recorded at baseline; medications and new use of glucose-lowering agents are monitored at every visit. FPG: fasting plasma glucose; HbA1c: glycated haemoglobin; CBC: complete blood count; HOMA-IR/β: Homeostatic Model Assessment for insulin resistance / β-cell function; EBIA: Brazilian Food Insecurity Scale; SISVAN: Brazilian Food and Nutrition Surveillance System; IPAQ-LF: International Physical Activity Questionnaire, long form; PSQI-BR: Pittsburgh Sleep Quality Index, Brazilian version; PSS-10: Perceived Stress Scale-10; SF-36: Short Form-36; MAAS: Mindful Attention Awareness Scale; ABEP: Brazilian Economic Classification Criteria.

### Sample Size Calculation

The sample size was calculated to detect differences in the cumulative incidence of T2D at 36 months between each intervention arm (PROVEN-DIA and TelePROVEN-DIA) and the control group. Based on evidence from systematic reviews [11], we assumed a cumulative incidence of 0.13 (13%) in the intervention arms and 0.23 (23%) in the control arm. Using a two-sided Bonferroni-adjusted significance level of α = 0.025 (to account for two primary comparisons), 90% power, and a 1:1:1 allocation ratio, 348 participants per group are required. Accounting for an anticipated 20% loss to follow-up over 36 months, the target sample size is 1,305 participants (435 per group), rounded to ensure equal distribution across the 30 investigation sites. Sample size calculations were performed in R software.

### Outcome

The primary outcome is the cumulative incidence of T2D at 36 months, assessed at each follow-up visit (6, 12, 24, and 36 months). T2D is diagnosed when both fasting plasma glucose ≥126 mg/dL and HbA1c ≥6.5% are simultaneously confirmed at the same assessment visit [25], requiring two concordant tests to maximize diagnostic specificity and minimize misclassification in an asymptomatic population. The oral glucose tolerance test (OGTT) was not included as a routine diagnostic criterion, as fasting plasma glucose and HbA1c are recommended as first-line tests in Brazilian and international guidelines and are feasible within the Brazilian Unified Health System (SUS), where OGTT is not routinely available in primary care.

Secondary outcomes were selected based on Brazilian Diabetes Society and international guideline recommendations for cardiometabolic risk monitoring in prediabetes [25] and are assessed at 6, 12, 24, and 36 months. They include: (1) Glycemic and metabolic parameters: fasting glucose, HbA1c, fasting insulin, HOMA-IR, HOMA- β, proportion of participants with controlled glycemia (fasting glucose <100 mg/dL and HbA1c <5.7%), and initiation of glucose-lowering medications; (2) Anthropometric measures: body weight, BMI, waist circumference; (3) Dietary quality and intake, and energy intake from ultra-processed foods; (4) Physical activity and sedentary behavior: minutes per week of moderate-to-vigorous physical activity (MVPA), proportion achieving ≥ 150 minutes/week of MVPA, and daily sedentary time; (5) Psychosocial and behavioral parameters: sleep quality, perceived stress, health-related quality of life, mindful attention awareness, alcohol consumption, and smoking status; (6) Additional metabolic parameters: lipid profile (total cholesterol, LDL-cholesterol, HDL-cholesterol, triglycerides), blood pressure, and renal function (creatinine, estimated glomerular filtration rate).

### Statistical Analysis

This is a parallel-group randomized controlled trial; treatment arm is treated as a between-subjects factor, while the repeated assessments at 6, 12, 24, and 36 months constitute a within-subjects (time) factor. Baseline characteristics will be described by study arm. Categorical variables will be summarized as absolute and relative frequencies, and continuous variables as means with standard deviations or medians with interquartile ranges, according to their distribution. Continuous outcomes with markedly skewed distributions (e.g., HOMA-IR, triglycerides) will be log-transformed prior to analysis to satisfy model assumptions and back-transformed for interpretation.

For the primary outcome (time to T2D diagnosis), Kaplan-Meier survival curves will be constructed for each study arm, and the cumulative incidence at 36 months will be estimated. Differences between groups will be assessed using the log-rank test. Hazard ratios (HRs) and 95% confidence intervals (CIs) will be estimated using Cox proportional hazards regression models, adjusted for baseline age, sex, and BMI. The proportional hazards assumption will be assessed using Schoenfeld residuals.

Longitudinal comparisons of secondary outcomes between study groups will be conducted using mixed-effects models, incorporating time, treatment group, and their interaction (group × time) as fixed effects, and participant as a random effect. The identity link function will be used for continuous outcomes, and the logit link function will be applied for binary outcomes. Effects will be expressed as mean differences for continuous outcomes and odds ratios (ORs) for binary outcomes, both with 95% confidence intervals. Adjustment for multiple comparisons will be performed using the Bonferroni method when applicable. For all regression models, underlying assumptions will be evaluated (including normality and homoscedasticity of residuals through diagnostic plots, linearity of continuous covariates, and multicollinearity assessed via variance inflation factors) with modeling decisions adjusted accordingly.

Analyses will follow the intention-to-treat principle, including all randomized participants in their assigned groups regardless of adherence to the intervention. For the primary outcome, participants without a diabetes diagnosis during follow-up will be censored at the time of last contact or end of study, as appropriate for time-to-event analysis. Prior to analysis, outliers and biologically implausible values will be screened using distributional checks and predefined plausibility ranges; primary analyses will be conducted on the complete dataset and repeated with identified outliers excluded as a sensitivity analysis. Missing data for continuous secondary outcomes will be handled using multiple imputation by chained equations (MICE), under the assumption of missing at random. Adjusted models will include investigation site as a random effect, and baseline age, sex, and BMI as fixed covariates.

Use of pharmacological co-interventions (including glucose-lowering agents, antihypertensives, lipid-lowering drugs, and psychotropic medications) initiated during follow-up will be systematically documented at each assessment visit and included as time-varying covariates in adjusted models to account for their potential influence on outcomes. In addition, a pre-specified subgroup analysis will examine whether the effect of the intervention differs according to initiation of pharmacological co-interventions during follow-up, and interaction terms will be included in regression models to formally test effect modification.

Pre-specified subgroup analyses will examine potential effect modification by sex, age, geographic region, baseline sedentary behavior, baseline consumption of ultra-processed foods (categorized by tertiles of percentage of total energy intake), baseline BMI category, baseline hypertension, baseline socioeconomic status, and baseline use of beta-blockers or thiazide diuretics. Interactions will be tested by including product terms in regression models. Sensitivity analyses using a per-protocol approach will be conducted for selected secondary outcomes, including body weight, fasting plasma glucose, and HbA1c. The per-protocol population will be defined as participants who attended ≥75% of scheduled intervention sessions. Additional sensitivity analyses will exclude participants who initiated glucose-lowering medications during follow-up.

All statistical analyses will be performed using R software (version 4.4.1 or higher; R Core Team, 2024). For the two primary comparisons (each intervention arm versus the control group), a two-sided Bonferroni-adjusted significance level of α = 0.025 will be applied, consistent with the sample size calculation. For secondary and exploratory analyses, a two-sided significance level of 0.05 will be used, with Bonferroni adjustment for multiple comparisons where applicable. No formal interim analyses for efficacy or futility are planned. The trial will continue until the planned sample size is reached and follow-up is complete, unless serious safety concerns emerge that warrant early termination. Safety data are reviewed continuously by the Coordinating Center and reported quarterly to local research ethics committees.

### Data Management Plan

Each investigation site is responsible for collecting data using standardized source documents and for transcribing them into electronic case report forms (eCRFs). Data are collected via two secure, web-based systems: REDCap (Research Electronic Data Capture) for screening, randomization, follow-up data, and statistical export [42]; and Vivanda®, a web-based system utilizing Brazilian food composition tables, for processing 24-hour dietary recalls collected on paper forms. Access is role-based and restricted to authorized personnel in accordance with Good Clinical Practice guidelines. Built-in validation rules flag inconsistent entries for immediate correction [43]. The Coordinating Center conducts monthly database reviews and issues data queries as needed. All data are stored on secure servers with automated backups and audit trails. The database will be locked after completion of follow-up and query resolution. All procedures comply with Brazilian data protection regulations, particularly Brazilian General Data Protection Law [44].

### Training and Monitoring

All investigation sites receive standardized training prior to study initiation, including an in-person workshop and standard operating procedures (SOP) manual. Training emphasizes standardized intervention delivery, behavioral counseling techniques, and data collection procedures. Training is differentiated by study arm assignments and role: specific materials are provided for Lifestyle Counselors and Lifestyle Coaches, while Control arm personnel receive training on standard lifestyle guidance delivery. Physical activity counseling receives particular attention in training, given the regional heterogeneity in infrastructure, access to exercise facilities, and population activity patterns across Brazil; standardized behavioral strategies and goal-setting frameworks are employed to ensure consistency in this domain regardless of local context. Ongoing refresher training and technical support are provided to maintain protocol fidelity throughout the study.

The Coordinating Center implements risk-based monitoring through remote assessments to ensure data quality, protocol compliance, and participant safety. Quarterly monitoring reports track recruitment progress, follow-up completion rates, data quality indicators, and adverse events, and are shared with investigation sites to support continuous quality improvement. Monthly follow-up meetings are held with site personnel to address questions and implement corrective actions as needed promptly. Corrective and preventive actions are promptly implemented when issues are identified.

### Safety Considerations

Given the lifestyle-based nature of the intervention, the study is classified as minimal risk. Adverse events (AEs) are monitored through active surveillance during scheduled interactions and spontaneous participant reports. All AEs are documented in accordance with protocol specifications. Serious adverse events (SAEs) are reported to the Coordinating Center and local research ethics committees in accordance with national regulatory requirements. Participants requiring medical care are referred to appropriate health services.

Given the minimal-risk nature of this lifestyle intervention, an independent Data Safety Monitoring Board (DSMB) was not deemed necessary. The Coordinating Center conducts safety monitoring through systematic adverse event surveillance, with annual safety reports reviewed by the study leadership team and local research ethics committees.

### Criteria for discontinuing or modifying allocated interventions

This is an intention-to-treat trial; all randomized participants will be included in the final analyses regardless of intervention adherence. Participants who discontinue the intervention will continue to be followed for outcome assessments unless they withdraw consent. Intervention may be discontinued due to participant request, pregnancy, medical conditions incompatible with the intervention, or SAEs. Participants who discontinue are encouraged to attend all scheduled follow-up visits. If a participant cannot be located after randomization, investigation sites will make multiple contact attempts. If the outcome cannot be determined after exhaustive efforts, the data will be treated as missing and handled using multiple imputation. Participants may be transferred between sites, when necessary, with data analyzed according to the original randomization site.

### Strategies to improve adherence to interventions

Participant retention is supported through personalized telephone calls and text messages sent by study staff prior to each scheduled visit, with rescheduling offered according to participant availability. Participants who miss a visit are actively followed by site staff. The motivational kit and phase-specific participation certificates serve as additional engagement incentives across all three arms.

### Relevant concomitant care permitted or prohibited during the trial

Participants are advised to avoid external individualized nutritional counseling or supervised physical activity programs during the study period. Routine medical care unrelated to structured lifestyle counseling remains permitted.

### Data Sharing Statement

De-identified individual participant data and relevant documentation will be available upon reasonable request after publication, subject to ethical approvals, for academic, non-commercial purposes.

### Dissemination

Upon completion, participants receive a personalized health report and are encouraged to maintain lifestyle changes. Those who develop T2D are referred for diabetes management. If intervention superiority is confirmed, control participants will be offered an effective program. Study findings will be disseminated through peer-reviewed publications, conferences, stakeholder engagement, ClinicalTrials.gov, and made available to participants upon request.

### Study Status and Timeline

Recruitment started in December 3rd 2024 and, as of June 2026, 944 participants have been enrolled. Recruitment is expected to be completed by the end of the third quarter of 2026. Each participant is followed for 36 months, with final visits anticipated by December 2029 and primary results by 2030.

## Discussion

The PROVEN-DIA trial addresses critical gaps in diabetes prevention research by translating efficacy findings from controlled settings to the heterogeneous Brazilian context, where geographic, socioeconomic, and healthcare system barriers pose substantial challenges to large-scale program implementation. Beyond evaluating effectiveness, the trial makes three methodological contributions that distinguish it from prior prevention trials. First, it provides the first large-scale evaluation of a structured diabetes prevention program across Brazil’s diverse, continental context [17,18]. Second, it directly compares two scalable delivery formats, hybrid (PROVEN-DIA) and telehealth-only (TelePROVEN-DIA), against usual care, addressing whether telehealth can achieve effectiveness comparable to hybrid delivery while overcoming geographic access barriers. Third, the intervention was deliberately designed for delivery by primary care professionals rather than specialists, reflecting the Brazilian Family Health Strategy structure, in which multidisciplinary teams provide comprehensive care with limited specialist access [45,46].

An important strength of PROVEN-DIA is its rigorous alignment with Brazilian public health policy: all intervention materials are based on the Brazilian Dietary Guidelines and the Brazilian Physical Activity Guidelines [26,27], facilitating potential adoption within the SUS if the program proves effective. The multicenter design spanning sites across all five Brazilian regions, ensures that the protocol reflects the country’s regional, cultural, and socioeconomic diversity. Prior to implementation, intervention protocols were validated through a modified Delphi technique involving a multidisciplinary expert panel, ensuring cultural appropriateness and feasibility [21]. This design was built on prior evidence from the research team: a multicenter pilot trial demonstrated operational feasibility and preliminary efficacy in improving dietary quality [22], and qualitative research identified key implementation barriers and facilitators [47]. These features collectively position PROVEN-DIA as a prevention strategy designed not only for scientific rigor but for real-world scalability within Brazil’s public health system, where the growing burden of diabetes reinforces the urgency of effective, guideline-based prevention programs [1,25,49].

Several limitations warrant acknowledgment. As a lifestyle intervention trial, blinding of participants and providers is not feasible. The multicenter scale may introduce implementation heterogeneity, reflecting variations in local infrastructure and protocol adaptation across sites. Behavioral outcomes rely partly on self-reported data including dietary intake and physical activity, which may be subject to recall and social desirability bias, though validated instruments were employed to mitigate this risk. Objective monitoring measures were not systematically incorporated; however, the combination of multiple validated self-report instruments across behavioral domains partially compensates for this constraint. The oral glucose tolerance test (OGTT) was not included as a routine diagnostic criterion for T2D ascertainment, which may result in missed cases identifiable by OGTT alone, potentially underestimating the true incidence of T2D and reducing internal validity. This decision reflects real-world diagnostic practice within the Brazilian public health system, where fasting plasma glucose and HbA1c are the recommended first-line tests, and is expected to enhance the external validity and scalability of the findings [25]. Participants may also initiate pharmacological treatments during follow-up including GLP-1 receptor agonists, SGLT2 inhibitors, metformin, or other glucose-lowering medications as part of routine clinical care; these will be systematically recorded to allow adjustment for potential confounding. Finally, the 36-month follow-up presents retention challenges inherent to long-term prevention trials, mitigated by structured adherence strategies built into the protocol.

Upon completion, this trial will provide the first rigorous evidence on diabetes prevention program effectiveness and telehealth scalability in Brazil. The study’s biorepository and multidimensional data collection enable future secondary analyses examining biological mechanisms, long-term outcomes, and cost-effectiveness, ultimately contributing to evidence-based primary care guidelines and health policies aimed at reducing the growing burden of diabetes in Brazil and similar contexts globally.

## Data Availability

No datasets were generated or analyzed during the current study. All relevant data from this study will be made available upon study completion

## Acknowledgments

The authors gratefully acknowledge Juliana de Carvalho Opipari, Rozana Mesquita Ciconelli, Rodrigo Quirino dos Reis, Mariana Beraldo da Costa Saad, and Dante Dianezi Gambardella for their valuable contributions to the development of the project and the acquisition of funding that supported this study.

## Supporting information

**S1 Checklist.** SPIRIT 2025 checklist.

**S2. File.** Eligibility Criteria for Participant Selection in the PROVEN-DIA Trial

**S3 Checklist.** TIDieR Checklist: Template for Intervention Description and Replication of the PROVEN-DIA Intervention

**S4. File.** S4 Intervention Framework, Components, and Implementation Strategy of PROVEN-DIA and TelePROVEN-DIA

**S5 File.** Usual care control group protocol.

